# COVID-19 PANDEMICS: HOW FAR ARE WE FROM HERD IMMUNITY?

**DOI:** 10.1101/2020.12.19.20248571

**Authors:** Carlos Hernandez-Suarez, Efren Murillo-Zamora, Francisco Espinoza Gómez

## Abstract

**Objectives:** to estimate the current number of total infections in a region in order to measure the progress of the epidemic with the purpose of reopening activities and planning the deployment of vaccines.

**Study design:** We recovered estimates of the basic reproductive number (R_0_) and the Infection Fatality Risk (IFR) as well as the number of confirmed cases and deaths in several countries.

**Methods:** this works presents an expression to estimate the number of remaining susceptible in a population using the observed number of SARS-CoV-2 related deaths and current estimates of R_0_ and IFR.

**Results:** the epidemic will infect most of the population causing 2.5 deaths per thousand inhabitants on average, and herd immunity will be achieved when the number of deaths per thousand inhabitants is close to two. This work introduces an expression to provide estimates of the number of remaining susceptible in a region using the reported number of deaths.

**Conclusions:** any region with fewer than 2.5 deaths per thousand individuals will continue accumulating deaths until this average is achieved, and the infection rate will exceed the removal rate until the number of deaths is about two deaths per thousand, when herd immunity is reached. Waves may occur in any region where the number of deaths is below the herd immunity level.

## INTRODUCTION

Although the COVID-19 pandemic possesses a relative low lethality rate, the actual burden of the disease thus far has been enormous. Many economic activities have had to be suspended or reduced and the pressure to reopen schools has been quite substantial. More information is required for policymakers and the general public to make informed decisions and to avoid potential civil unrest. The competition for developing a vaccine is intense, with about 321 vaccine candidates with 32 clinical trials currently in progress [1]. Besides the safety concerns [2,3] the additional economic cost for the purchase and deployment of vaccines has not been factored into the substantial burden of the disease. To date, three vaccines have ended phase III trials and appear to show promising efficacy: Moderna, Pfizer-BioNTech and AstraZeneca-Oxford.

At present, without available vaccines or effective pharmaceutical treatments, the decision on whether to reopen activities or not depends primarily on the number of additional projected infections and deaths caused because by opening schools, allowing public gatherings, opening tourist sites, increasing number of people allowed in cinemas, restaurants, buses, planes, etc. For a highly infectious virus like SARS-CoV-2, any change in policy should be based on the remaining susceptible population in the affected region, that is, in the progress of the disease.

Using the number of individuals accessing private or public hospitals as a surrogate of the current number of infections, or even the number of confirmed cases, is not very accurate. This is the case because numbers strongly depend on the availability of medical services that are not always accessible to large segments of the population in many countries, either because of a lack of resources for test deployment or because of policies that disregard testing. The number of deaths is a more accurate indicator. If one calculates the IFR, that is, average number of infections that result in the death of one individual, this number can be used to estimate the number of infections that were required to observe the current number of deaths and the share of susceptible individuals in a well-defined population. Even though deaths are not reported as COVID-19 related, records may exist that shed light on the likelihood that the deaths were or were not caused by SARS-CoV-2. This strategy forms part of the policy adopted by some countries like Belgium, which is, without doubt, one of the reasons why this country reports one of the highest number of deaths per capita in Europe.

In order to estimate the progress of the epidemic in a region of interest we need to estimate two quantities: how many individuals will be infected (X) and how many infected there are at the moment (Y). The quotient Y/X is an estimate of the progress of the disease. In here we attempt to provide estimates of both quantities.

## METHODOLOGY

Two facts form the basis of this work. The first proposes that, with few exceptions, it is almost impossible to stop this pandemic by mitigation and control measures, and that the most that can be done is to reduce the infection rate (at a huge economic and social cost), which is commonly referred to as *flattening the curve*. These measures may be implemented effectively for some time, but it is extremely difficult to maintain due to the enormous social and economic costs. The second establishes that there are now several reliable estimates of the IFR available, from different studies. It is the confluence of these two facts that permits us to establish a surrogate for the progression of the epidemic in a region and calculate the share of susceptible available, which can be used as the basis to reopen activities and deploy vaccines.

### The expected number of SARS-CoV-2 infections

The basic reproductive number (R_0_) of a disease is the potential of disease transmission in the absence of any control or mitigation measures, and it is this potential that comes into play as soon as control or mitigation measures are suspended. Several studies have shown that the basic reproductive number R_0_ may be as high as 5.7 [4,5,6,7,8]. Here, we use the Susceptible-Infected-Removed (SIR) model introduced by Kermack and McKendrick [9] For this model, the estimated fraction of final, infected *f*, is approximately:

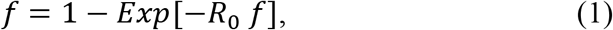

(for a probabilistic derivation, see [10]). For instance, if R_0_ = 5.7 the fraction of infected will be *f* = 0.99. However, a reduction of R_0_ by half will make *f* = 0.93, that is, it does not result in a very large difference in the size of the epidemic. However, reducing R_0_ by half is an incredibly difficult task that, in the absence of vaccines, involves mainly lockdowns and the use of face masks, which has a huge social and economic cost that cannot be sustained for long periods, except for rich countries or countries whose political or cultural organization allows their implementation. Nevertheless, even if a country manages to implement actions to reduce R_0_ to a value of less than one and can support citizens economically to maintain the lockdown for long periods, a handful of infected individuals who enter the country from abroad can restart the infection process if lockdowns are lifted, resulting in repeated waves (see Fig. 1 for examples in Belgium, France, Italy, Spain and Sweden). This is particularly true for a disease where the number of asymptomatic individuals is by far larger than that the number of symptomatic, making it difficult to control. For instance, the probability that *n* infected individuals will cause an epidemic is approximately 1 − (1/*R*_0_)^*n*^, which, for R_0_=5.7 and *n*=1 is about 0.82. Nevertheless, if we start with just 3 infectious, the probability grows to 0.99, this is the reason why it is unlikely that lockdowns will be effective to control the spread of the virus. As a consequence, Equation 1 can be seen as a quota of infections that must be fulfilled, as long as the fraction of remaining susceptible is larger than *f*.

**Fig 1:**
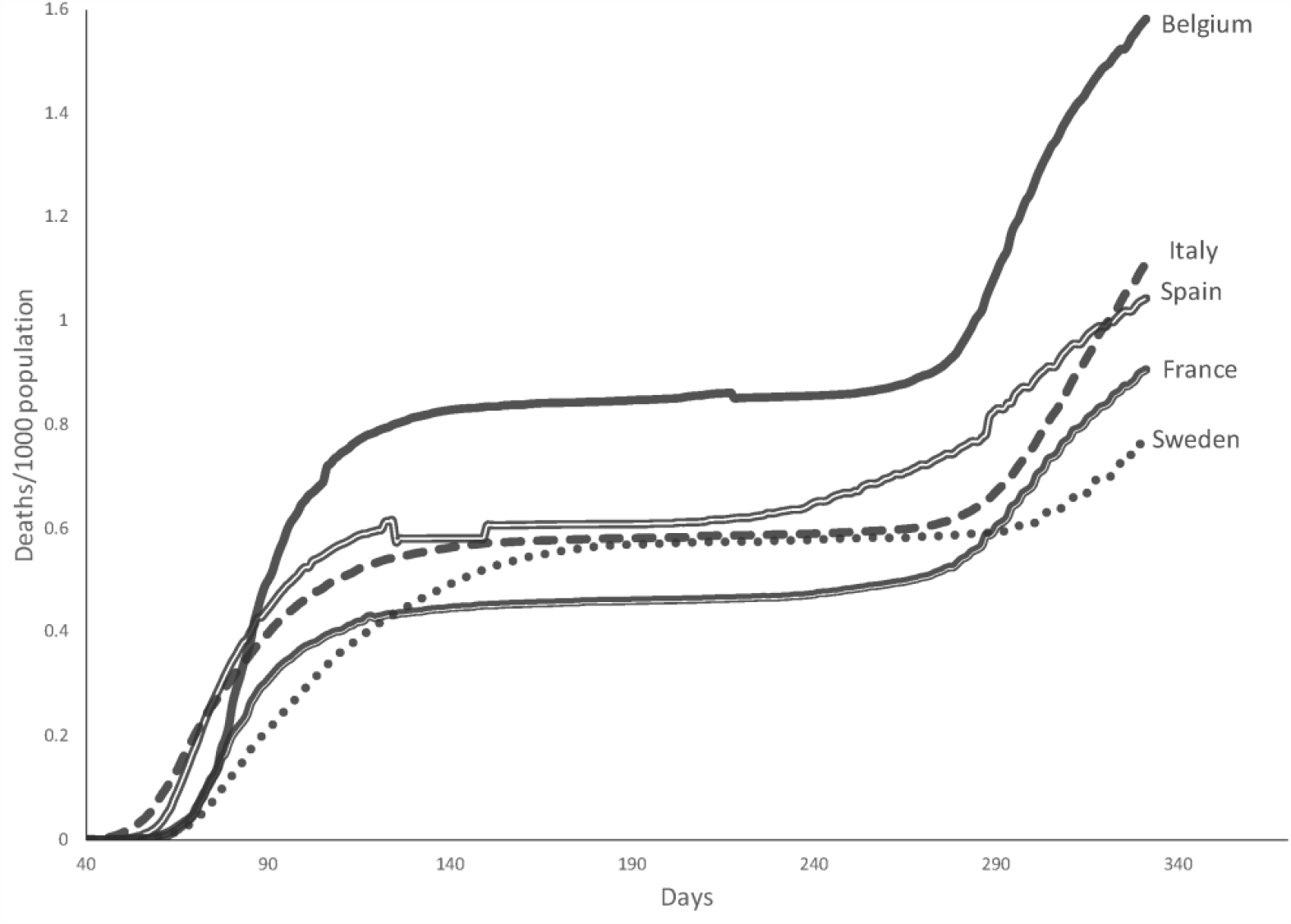
The appearance of a second wave. Accumulated number of deaths per thousand population. The use of fatalities instead of confirmed cases delays the detection of a wave but is more efficient in the sense that it does not depend on testing policies, availability or panic. The current number of deaths per thousand inhabitants for is Belgium: 1.58; France: 0.91; Italy: 1.11; Spain: 1.04; Sweden: 0.789. Data updated to 12/18/2020 from [11])

### The lethality of SARS-COV-2

In here we use the notation ‘IFR’ to indicate the true overall Infection Fatality Risk Fatality of SARS-CoV-2, given in expected deaths per thousand infections, whereas we use ‘IFR*’ to indicate the observed number of deaths per thousand individuals in a region of interest. Some attempts have been made to estimate IFR which would be a remarkable surrogate to measure the progress of epidemics in an arbitrarily defined region or population. However, the main drawback is that although the number of deaths from SARS-CoV-2 can be approximated, the total number of infected from the disease is elusive, especially considering that a large fraction of individuals that are infected are asymptomatic. Besides, the efficacy of immunity tests to detect who has been infected and recovered has been challenged with the findings that the ability to detect antibodies is reduced in a few days, especially in those with mild or no symptoms [12,13]. Recently, Eyre et al. [14] reported that a high fraction of individuals with none or mild symptoms may be undiagnosed mainly due to the calibration strategies of some standardized tests and concluded that samples from individuals with mild/asymptomatic infection should be included in SARS-CoV-2 immunoassay evaluations.

Some studies to estimate the IFR have been recently released for Iceland [15], India [16], Germany [17] and Denmark [18] among others. A median of 0.25% is reported in a review of 23 studies where the IFR was estimated by Ioannidis [19] which is about 2.5 deaths for each 1000 individuals. The study in Iceland, [15], the most comprehensive to date, proposes an IFR of 3 per thousand (95% CI, 2 to 6). This estimate deserves some caution since Iceland has a population of 322 thousand inhabitants with only 29 deaths when the estimate was calculated. In other studies that used transmission models fed with serological data, the IFR estimates were as high as 2.5-5 [20] and 3-8 [21 per thousand infections.

## RESULTS

Due to the high R_0_ of SARS-CoV-2 we conclude that most of individuals will be infected and the degree of advance of the epidemics can be estimated by measuring how far we are from the expected share of deaths, that is, using IFR*/ IFR. Thus, one can estimate the fraction of total infected at time *t*, asymptomatic or not, with:

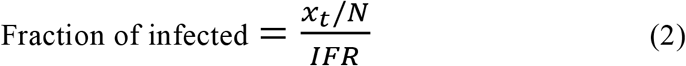

where *x*_*t*_ is the number of observed deaths at time *t* and *N* is the population size. The estimated fraction of susceptible as a function of the IFR in New Jersey and New York is shown in Fig. 2. We can see that if the IFR=2.5/1000, the fraction of remaining susceptible in New Jersey and New York is 18% and 26%, respectively.

**Fig 2:**
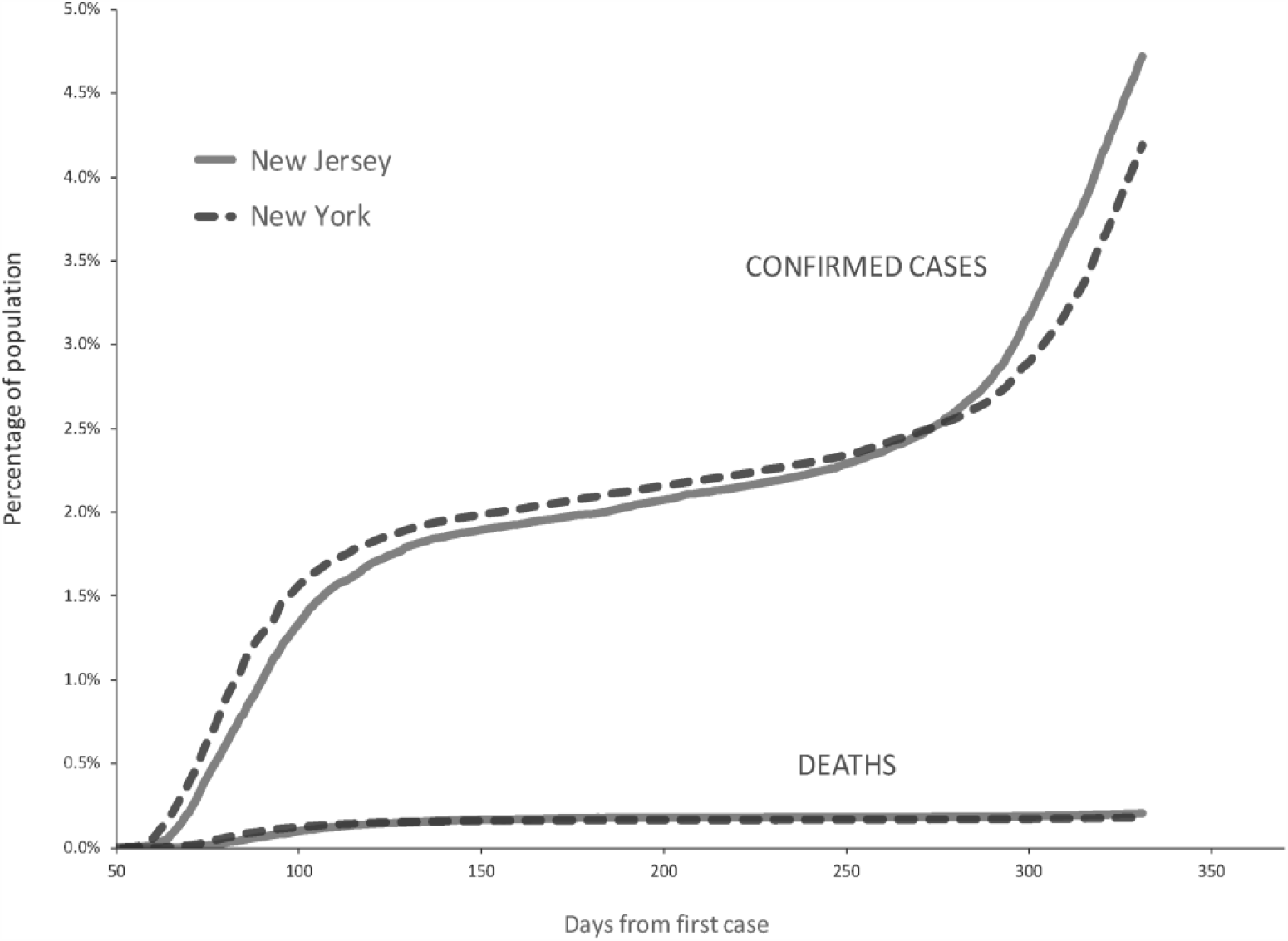
The fraction of remaining susceptible in the states of New Jersey and New York as a function of the lethality of SARS-CoV-2.

### Herd immunity

Herd immunity is achieved when the rate at which new infections are acquired equals the rate at which infected die or recover, which in an SIR occurs when the fraction of infected is 1 − 1/R_0_. Since a fraction IFR of these infected die, herd immunity occurs when the fraction of deaths equals IFR(1 − 1/R_0_). Using a conservative estimate of R_0_ = 5 and IFR = 2.5 per thousand infected, we conclude herd immunity occurs when the share of infected is 80%, that is, when the number of deaths equals 2 individuals per thousand. It is important to stress that herd immunity does not mark the end of the epidemic, but only the point at which recoveries are more likely than new infections.

## DISCUSSION

Using the IFR and the current number of deaths seems to be useful to estimate the progress of the disease and every effort has to be made to improve current estimates of the IFR. Hernandez-Suarez et al. [22] suggested that because of the high contagiousness of SARS-CoV-2, secondary cases resulting in fatalities in a household with at least one confirmed infected individual may be useful to estimate the IFR. This approach is significant because it would provide a large amount of data and require minimal testing.

Antibody testing in New York shows to date (12/12/2020) that out of 3 007 758 tests, 731 512 were positive for antibodies [23], that is, 24% of the population may be immune. This contradicts our findings that for an IFR as high as 2.5 per thousand the percentage of infected should already be close to 75% (see Fig. 3). Nevertheless, the fraction of people with antibodies is not a good surrogate of the fraction of infected individuals with SARS-CoV-2 because it underestimates the number of infected for two reasons: first, antibody testing is voluntary and it is natural to expect individuals with no symptoms to be less compelled to get tested than those who are symptomatic, resulting in a large percentage of infected not being tested, and second, some failures in testing have been reported, apparently due to calibration procedures that tend to fail in individuals with none or mild symptoms [14].

**Fig 3:**
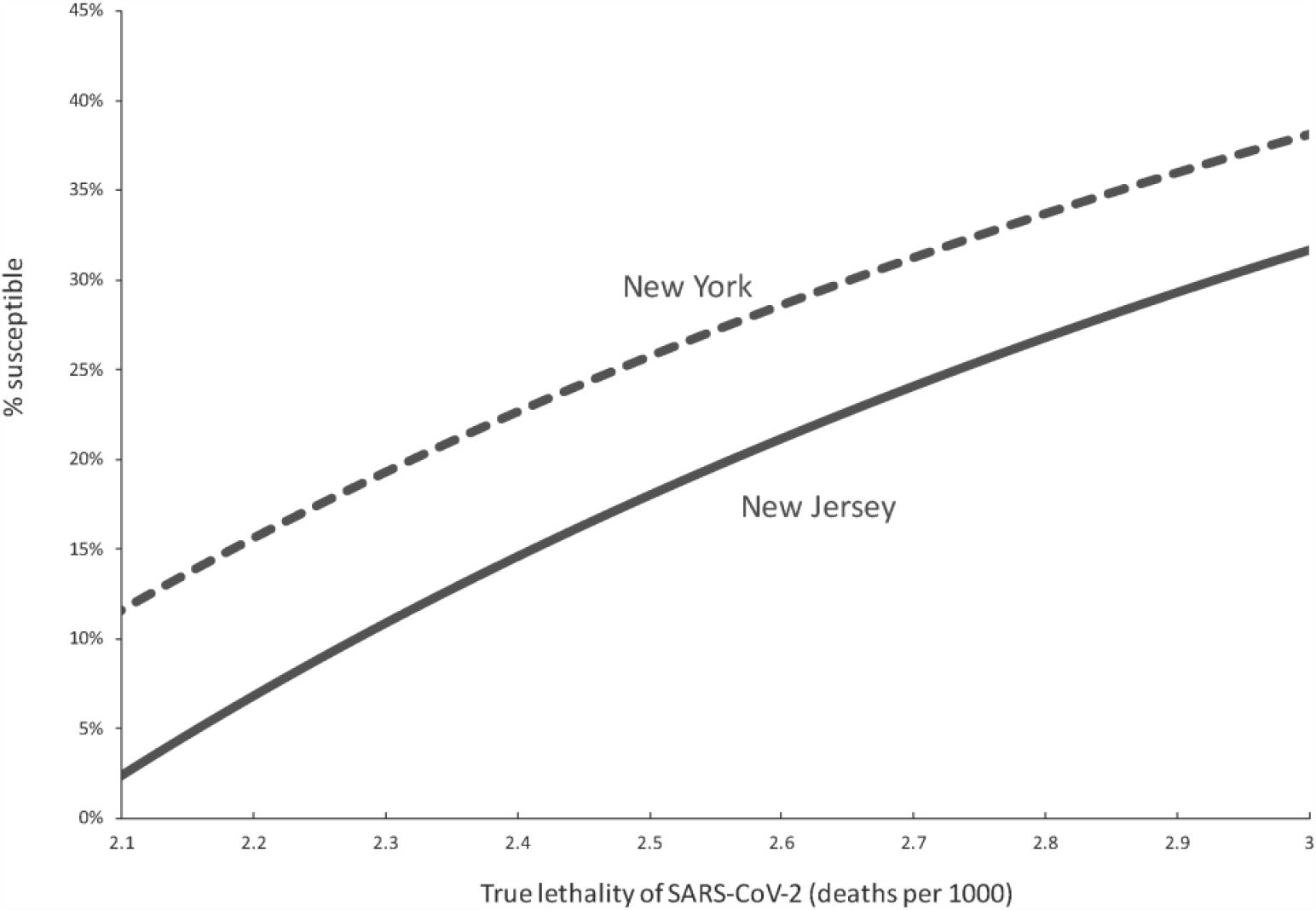
Percentage of the population that has been confirmed or died from SARS-CoV-2 in the states of New Jersey and New York. While the share of confirmed cases exhibits a second wave, that of deaths has remained steady. The current number of deaths per thousand inhabitants is: New Jersey: 2.049; New York: 1.856. Data updated to 12/18/2020 from [11].

The use of deaths as a more reliable surrogate of the number of infected is illustrated in Fig. 3. The current IFR* calculated for the states of New Jersey and New York is 2.049 and 1.856, respectively. In the las two months from 10/18/2020 to 12/18/2020, the percentage of the state of New Jersey’s population confirmed with SARS-CoV-2 and that of deaths increased 2.25% and 0.021%, respectively, that is, confirmed cases grew 107 more times than deaths. For the state of New York, the increase was 1.7% and 0.013% respectively, that is, 133 times more confirmed than deaths. Any attempt to attribute this uneven growth between confirmed and deaths to improvements in medical treatment, would be unreasonable. It is more rational to assume that the interest in being tested has increased.

The idea that the share of susceptible in New Jersey is relatively small depicted in Fig. 2 is supported by the number of active cases reported to date in the state (12/18/2020) is about 200 000. Unless they are fully quarantined, the infectious pressure from these individuals and those unreported must be huge, however, no new significant peak in deaths has been observed (see Fig. 3). The same happens in New York, with 371 000 active cases to date, although the share of susceptible is higher.

It is important to add that the uncertainty of how far the state is from achieving herd immunity is mainly driven by the uncertainty in the IFR estimate. Every effort should be made to obtain more accurate estimates.

The possibility of achieving herd immunity requires the existence of immunity after infection. New research indicates the presence of immunity for at least 6 months after infection [24,25,26]. In this study, spike IgG was relatively stable over 6 months after infection although CD4+ T cells and CD8+ T cells declined with a half-life of 3-5 months. This conveys good news for those regions well advanced in the epidemics that will soon achieve herd immunity and for those waiting for a vaccine.

Manaus, in Brazil, is a city where the COVID-19 pandemics has hit hard, with an estimated 76% attack rate in October based in an analysis of antibodies in a sample of blood donors [27]. To date (12/18/2020), there has been 3 167 confirmed deaths of COVID-19 [28]. The estimated population in Manaus is over 2 million, thus, using expression (2) we estimate that to date the share of infected in Manaus is 63%, which is our estimate of the progress of the epidemic so far in the city.

If we believe that only a fraction *p* of the COVID-19 related deaths was recorded, then expression (2) can be modified to:

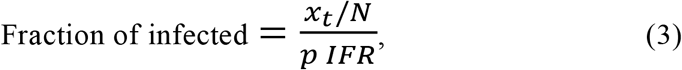

to obtain an improved estimate of the progress of the disease. For instance, if we believe 15% of deaths was unrecorded in Manaus, p=0.85 and this would increase our previous estimate of 63% infected to 74%, much closer to the estimated value in October [27].

Everything leads to the following question: what will be the purpose of a vaccine in a region where the number of deaths per thousand is already large enough to suggest there is only a small fraction of susceptible with limited chance of infection? This question is especially valid from the comprehensive study in Iceland that strongly suggests the existence of immunity due to infection [15]. Unless it is proven that immunity wanes beyond a protective level after some time, everything seems to indicate that those regions where the share of estimated susceptible is already low should have less priority in the distribution of vaccines. If these considerations are not taken into account, a vaccination campaign in a region where the proportion of deaths among the population is close to 2 will just give the impression of being effective but will not play a significant role, regardless of the efficacy of the vaccine.

## Data Availability

All data used is in public records and referenced

## Ethical approval

Not required: There is no data resulting from experimentation or sensitive whatsoever. All data used has been published elsewhere and the source has been properly referenced.

